# Management of the Covid-19 Health Crisis: A Survey in Swiss Hospital Pharmacies

**DOI:** 10.1101/2020.12.08.20237339

**Authors:** Laurence Schumacher, Yassine Dhif, Pascal Bonnabry, Nicolas Widmer

## Abstract

**Background:** The COVID-19 pandemic has put a lot of strain on health systems since 2020. A review of the Swiss hospital pharmacies responses during the first wave was performed to improve the quality of the pharmaceutical management of future health crises.

**Methods:** An electronic survey was sent to all head of hospital pharmacies in Switzerland. The questionnaire was organized into eleven clusters of questions and covered many topics regarding the management of the first wave of COVID-19. Data collection was conducted from May to June, 2020.

**Results:** Analyses were performed with 43 responses (66%) out of 65 questionnaires sent (at least one answer per questionnaire). 41% (17/41) of pharmacies had existing standard operating procedures or pandemic plans and 95% of them (39/41) created a steering committee to manage the crisis. 67% (29/43) created new activities to respond to the specific needs of the crisis. 67% (26/39) created new drug lists for: COVID-19-specific treatments (85% of pharmacies; 22/26), sedatives (81%; 21/26), anaesthetics (77%; 20/26) and antibiotics (73%; 19/26). Drug availability in COVID-19 wards was managed by increasing existing stocks (54% of pharmacies; 22/41) and creating extra storage space (51%; 21/41). Two drugs generated the most concern about shortages: propofol (49% of pharmacies; 19/39) and midazolam (44%; 17/39). Remdesivir stocks even ran out in 26% of pharmacies (10/39). Specific new documents were drafted to respond to medical needs with regards to drug administration (28% of pharmacies; 12/43), drug preparation (28%; 12/43) and treatment choices (23%; 10/43).

**Conclusions:** Swiss hospital pharmacies encountered many challenges related to the COVID-19 crisis and had to find solutions quickly, effectively and safely. The survey highlights the key role played by Hospital Pharmacy’s in many aspects during the pandemic by providing logistical and clinical support to medical and care teams. The lessons and experiences outlined could be used to improve the quality of the preparation for similar future events.

**KEY POINTS:**

▸ The COVID-19 pandemic generated unprecedented global demand for specific drugs, hand sanitizer solution, and other therapeutic products, particularly in critical care settings, highlighting the essential role of hospital pharmacists in such crises.
▸ Key COVID-19 responses at the hospital pharmacy level included staff flexibility with regards to changes in roles and procedures, communication, teamwork and solidarity, and the need to prepare business continuity plans and management dashboards ▸ Managing and facing complex pandemic response reveals the importance of involving hospital pharmacists in pandemic response steering committees at many levels. The lived experiences during the pandemic could have been reviewed and evaluated to raise awareness and guide future policy responses for when the next crisis occurs.

## BACKGROUND

The year 2020 was marked by the Coronavirus Disease 2019 (COVID-19) pandemic, which will certainly go down in history. In December 2019, the city of Wuhan, capital of Hubei Province in China, reported the first cases of pneumonia due to a new, previously unknown coronavirus(1). In February 2020, the World Health Organisation (WHO) officially named this new virus SARS-CoV-2 and the disease it caused COVID-19, short for Coronavirus Disease 2019. Human-to-human transmission has been confirmed(2).

The first case of COVID-19 in Switzerland was confirmed on 25 February 2020 in the canton of Ticino. On 28 February, in accordance with the Federal Act on Epidemics (EpidA), the Swiss Federal Council declared a “special situation” by presenting the COVID-19 Ordinance, and it prohibited gatherings of more than 1,000 people. At the beginning of March 2020, the number of cases detected became increasingly important in Europe, mainly in Italy, where the European outbreak started(3). On 11 March, the WHO declared the COVID-19 epidemic to be a pandemic. On 12 March, Switzerland had the third highest prevalence of COVID-19 of any European country affected by the coronavirus. On 16 March, the Swiss Federal Council declared an “extraordinary situation” (Art. 7 EpidA) and updated Ordinance 2 with new measures: the closure of non-essential businesses (including restaurants and leisure facilities) and partial border closures(4). Progressively, it also mobilised several conscript formations of the Swiss Armed Forces to assist the cantons with healthcare, logistics and security(5). The Swiss healthcare system did not reach saturation despite Switzerland being one of the most highly affected countries in the world, with prevalence increasing from 7.2 to 357 cases per 100,000 inhabitants between 9 March and 19 May 2020(6). As of summer 2022, Switzerland is facing already his seventh wave of the COVID-19 pandemic, resulting in a total of 55’396 hospitalizations and 13’436 deaths due to a COVID-19 infection since the beginning of the pandemic.

The services of hospital pharmacies were in high demand especially during this first wave of the pandemic because of their critical responsibility for the supply of therapeutic products. However, in the context of disasters, pharmacists were often forgotten or disregarded as a necessary disaster health care team member. In fact, pharmacist’s roles in disaster did not become of significant interest in the literature until the events of September 11, 2001(7). Prior to 2001, accepted roles for pharmacists generally focused on their contributions to logistics and supply chain management(8). In addition to this established role in supply chain logistics, several roles for pharmacists in disaster health management in general(9, 10) and pandemic(11) in particular have been identified mainly following disasters such Hurricane Katrina in 2005(12) or bushfire disaster in Australia in 2019(13).

Since the beginning of the COVID-19 pandemic, many studies have been published on the public health response, but few focus specifically on hospital pharmacy response and the role of pharmacists(14, 15, 16, 17, 18). Some described however already the response in Europe but with not so many details.(19, 20) All publications reporting in detail on the experience during the pandemic are essential to improve the quality of the preparation for such an event in the future.

The present study’s objective was thus to obtain an overview of the actions undertaken by hospital pharmacies across Switzerland as they responded to the challenges encountered during the first wave of the COVID-19 pandemic and to provide information and attitudes to improve the quality of the pharmaceutical management in future health crises.

## METHOD

A closed survey using a web-based open questionnaire, multicentre, designed on the SurveyMonkey^®^ platform (SurveyMonkey, San Mateo, CA, USA) was conducted. Various questions in the survey emerged from initial feedbacks from fellow hospital pharmacists. Several practical problems experienced daily have made it possible to suggest the challenges faced by hospital pharmacists during the COVID-19 crisis.

All the questions were organised into eleven sections: general information about the respondent, the hospital pharmacies and the hospital, management of this crisis, human resources management, drugs used primarily in intensive care units (ICUs), drugs used specifically for treating SARS-CoV-2, drug management in COVID-19 care units (including ICUs), hygiene, support for medical and care teams, care management for patients recovering from COVID-19, other problems encountered, and future perspectives. Altogether, it consisted of 67 questions and used adaptive design to create a custom pathway through the survey depending on the respondent’s answers. We used conditional branching to create a custom path through the survey, depending on the respondent’s answer. Therefore, not all participants needed to answer every question. Each question can therefore have a different number of respondents. To manage the non-response error and these “missing data”, each question uses its own denominator. Most of the questions were multiple-choice and allowed multiple answers. All questions are available in appendix I.

The questions were reviewed and validated by the steering committee of the Swiss Society of Public Health Administration and Hospital Pharmacists (GSASA) and the survey was tested on the web platform by two head pharmacists (GSASA members) before fielding of all other members. The large majority of acute hospital pharmacists in Switzerland are members of GSASA.

Data were collected between 19 May and 19 June 2020 via an anonymous questionnaire sent out by individually by email (to protect unauthorized access) to all the country’s head hospital pharmacists affiliated to the GSASA (corresponding at 65 people; inclusion criteria), to avoid duplicate entries (to prevent “multiple participation” of participants). Email reminders were sent out at two, three and four weeks after the first mailing. Data were analysed using standard descriptive statistics. Means and proportions were calculated using the total number of participants responding to a given question. Responses left blank were coded as missing data and handled using a list wise deletion method. The respondents who answered only general information were excluded.

The research protocol, which did not imply patient data collection, has been presented to the Cantonal Research Ethics Committee Geneva, which waived an ethical oversight.

## RESULTS

### GENERAL INFORMATION

Responses of 43 participants were obtained out of 65 surveys sent, which represented a participation rate of 66%. The number of answers for each question are available on the appendix The participants completed the survey in about 39 min. in average. Switzerland’s population is spread across four linguistic regions: German-speaking(G), French-speaking(F), Italian-speaking(I) and Romansh-speaking(R). Most of our study participants worked in German-speaking regions (79%; 34/43) and only one of 43 participants (2%) worked in each of the Romansh-speaking and Italian-speaking regions. The remaining participants worked in French-speaking regions (19%; 8/43). To simplify the analysis, French- and Italian-speaking participants were grouped together(F/I), and so were German- and Romansh-speaking head hospital pharmacists(G/R).

### MANAGEMENT OF THE CRISIS

During the first wave of the COVID-19 outbreak, 41% (17/41) of hospital pharmacies had previously prepared internal standard operating procedures regarding disaster management plans whether general disaster Standard operating procedures (17%; 7/41) or specific pandemic plans (24%; 10/41); 18% (7/39) of hospital pharmacies had a business continuity plan prior to the pandemic (38% F/I, 3/8; 13% G/R, 4/31); and an equal number of hospital pharmacies (18%; 7/39) created a plan especially to manage the pandemic. Of those hospital pharmacies which had a pandemic/disaster management plan prior to the COVID-19 crisis, 56% (9/16) did however not a business continuity plan have. This percentage rose to 70% (16/23) among hospital pharmacies which did not have a pandemic/crisis management plan prior to the COVID-19 crisis. The details of the business continuity plans are listed in Table 1.

**Table 1.**
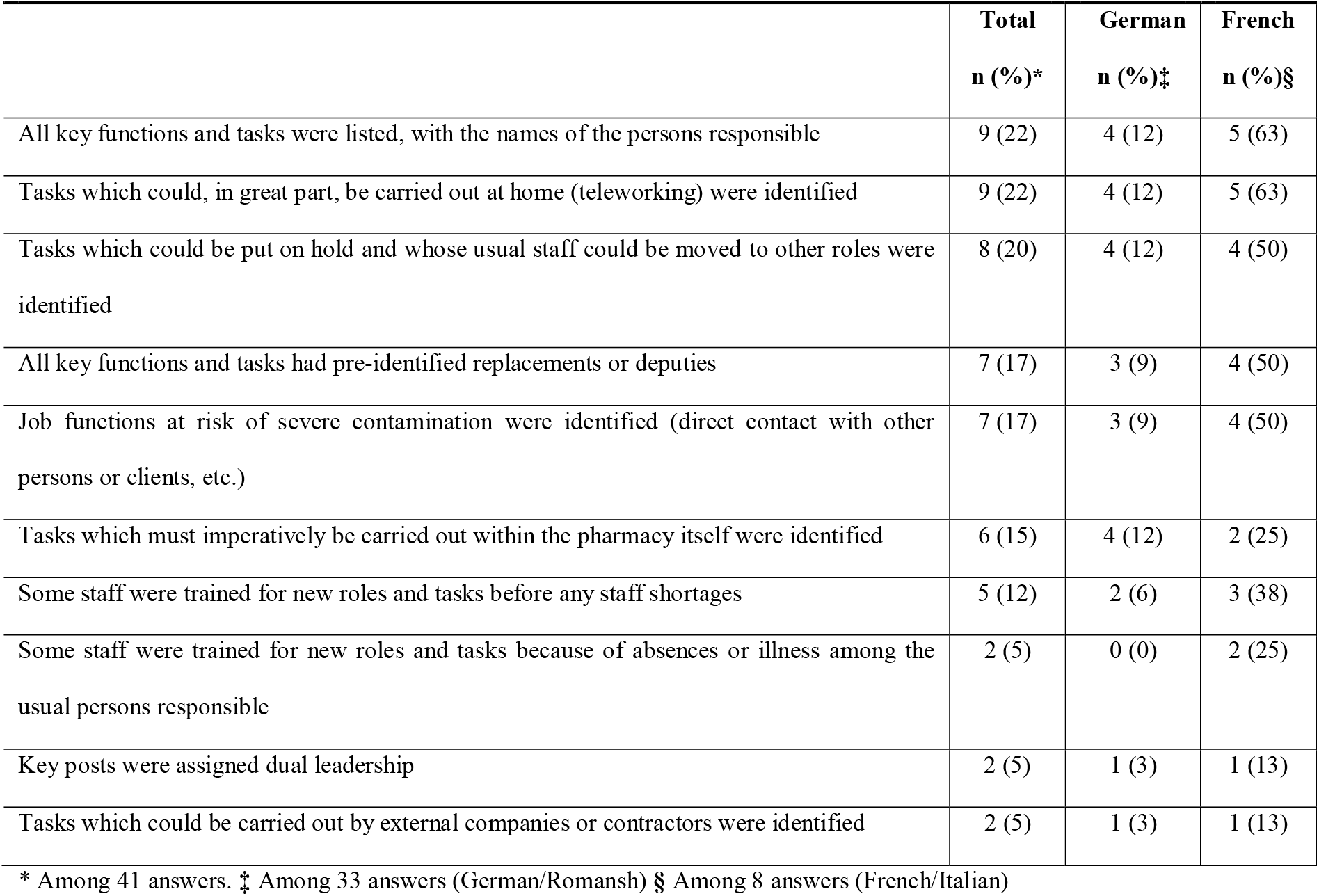
Business continuity plan.

Most hospital pharmacies (95%; 39/41) created a steering committee to manage the crisis. This was often composed of the head pharmacist (61%; 25/41), a member of the pharmaceutical logistics unit (34%; 14/41), a representative from outside the pharmacy (32%; 13/41), a representative of the hospital’s general crisis management team (27%; 11/41), a representative from the cantonal authorities (22%; 9/41) and a member of the clinical pharmacy unit or the pharmaceutical assistance unit (20%; 8/41). Among hospital pharmacies with a disaster management plan, 53% (9/17) created paper or electronic dashboards or another management tool (i.e. an Excel^®^ file) especially for the COVID-19 crisis, but 24% (4/17) had no dashboards at all. The remaining 24% (4/17) used previously prepared dashboards. Of the hospital pharmacies without prior Standard operating procedures, 61% (14/23) created dashboards specifically to manage the crisis. The panels of those dashboards are listed in Table 2. It is worth noting that only two hospital pharmacies (5%; 2/43) had a panel of risks management.

**Table 2.**
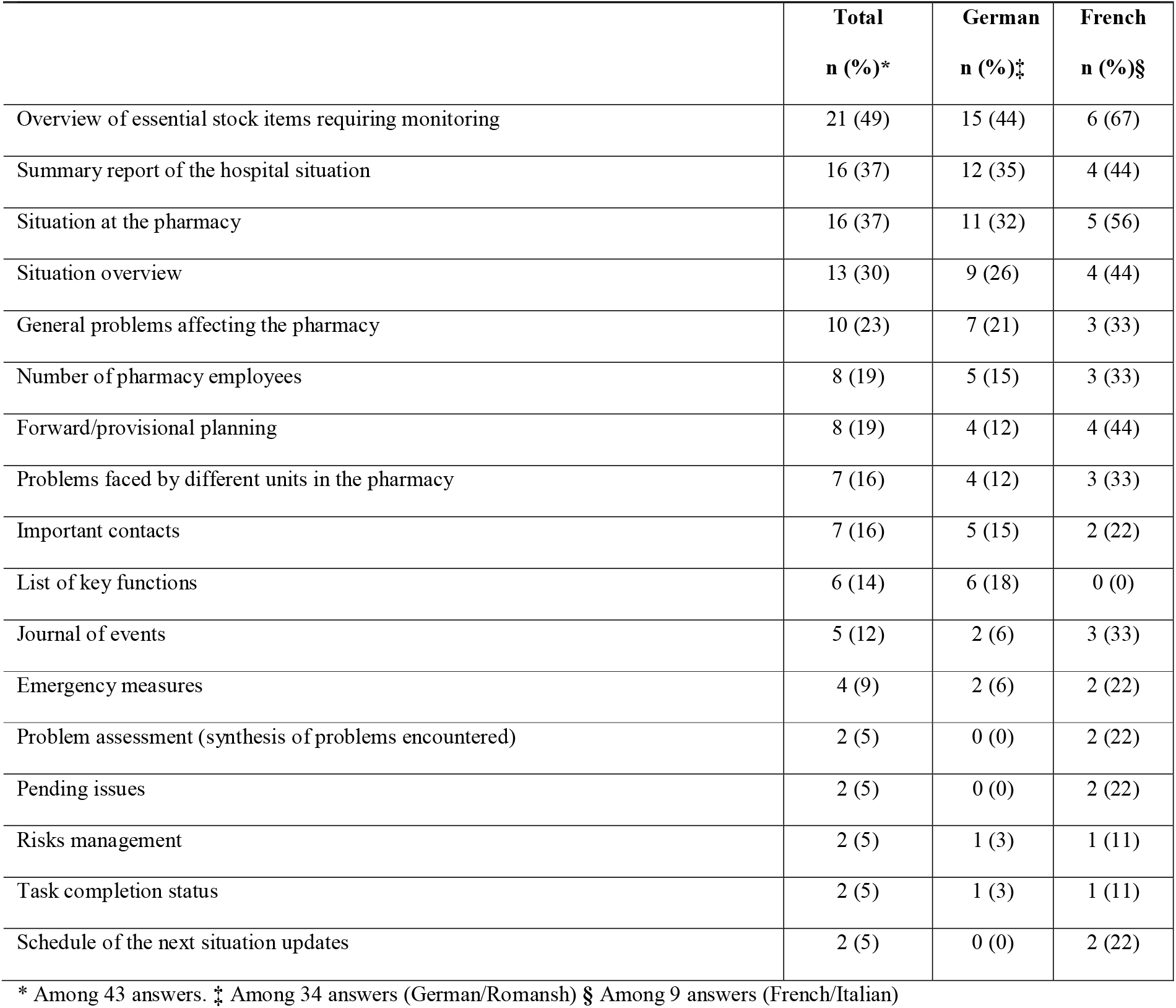
Information present on dashboard panels for the management of the COVID-19 crisis.

### HUMAN RESOURCES MANAGEMENT

Hospital guidelines were the main source of human resource management advice during the COVID-19 outbreak (70%, 30/43). The main changes in human resource management carried out are summarised in Table 3. Role changes occurred in the following fields: disinfectants manufacturing (collecting bottles, production, and filling done by pharmacists and volunteers), administrative work for ICUs (done by pharmacy technicians), pharmacists also acted as pharmacy technicians on wards and logisticians in drug distribution, with 9% (4/43) of hospital pharmacies reassigning pharmacists to these tasks.

**Table 3.**
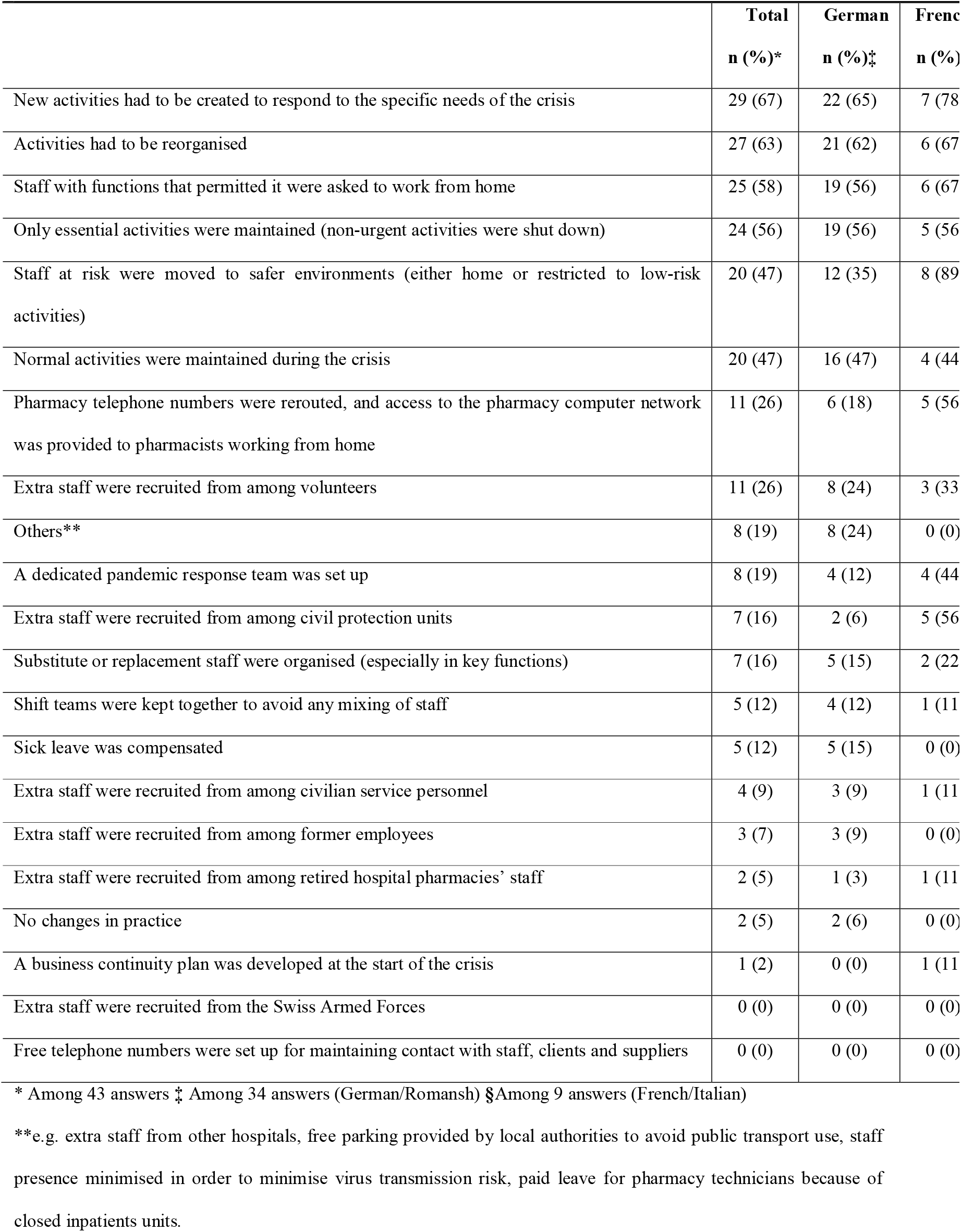
Human resources management.

### DRUGS MANAGEMENT

To limit risks of any shortages, 56% of hospital pharmacies (23/41) had anticipated reserve supplies (100% F/I, 8/8; 45% G/R, 15/33). A list of drugs used for treating COVID-19 patients and their supply problems is compiled in Figure 1. 34% of hospital pharmacies (14/41) imported drugs from the European Union (63% F/I, 5/8; 27% G/R, 9/33), but only 7% (3/41) (25% F/I, 2/8; 3% G/R, 1/33) had to import drugs from a country outside those usually used. 10% (4/41) of hospital pharmacies asked a statistician to provide stock forecasts (13% F/I, 1/8; 9% G/R, 3/33). Less than a quarter of hospital pharmacies (22%; 9/41) managed never to run out of drug stocks at all (13% F/I, 1/8; 24% G/R, 8/33). Drug stocks on wards treating COVID-19 patients were sometimes managed by assigning dedicated pharmacy technicians (29%; 12/41). 76% of hospital pharmacies (31/41) managed the risk of shortages by strictly monitoring the drugs dedicated to COVID-19 patients (63% F/I, 5/8; 79% G/R, 26/33). 63% of hospital pharmacies (26/41; 75% F/I, 6/8; 61% G/R, 20/33) found alternative drugs and proposed these instead to medical staff or their healthcare institution. Alternatives were proposed to guarantee the continuity of care and save the stocks. 27% of hospital pharmacies (11/41) had to prepare alternative protocols in partnership with medical or care staff in case these potential shortages occurred (50% F/I regions, 4/8; 21% G/R regions, 7/33). 51% of hospital pharmacies (20/39) created specific drug lists for care units treating COVID-19 patients (75% F/I, 6/8; 65% G/R, 20/31). Types of drugs contained in these lists are summarised in Table 4.

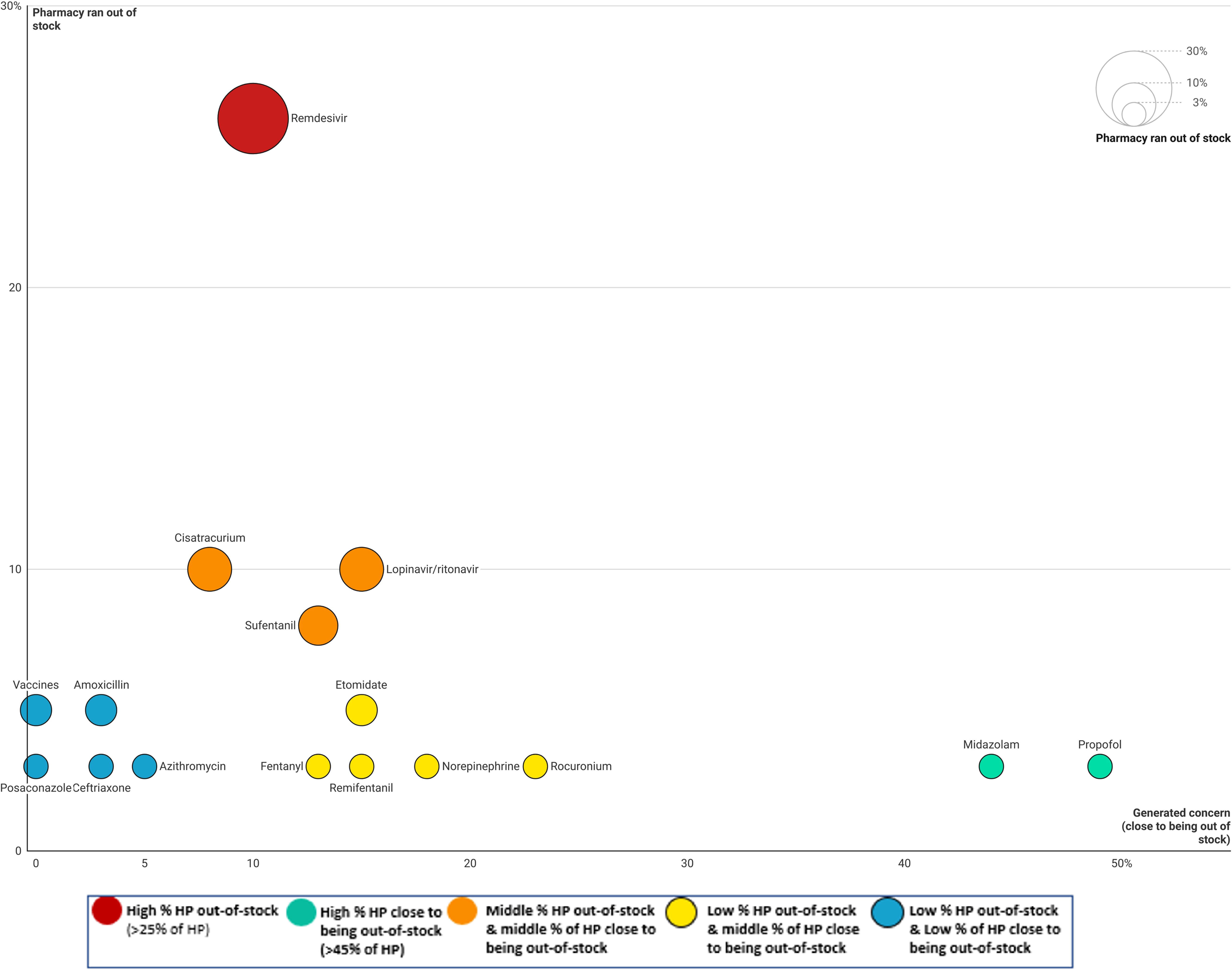

**Table 4.**
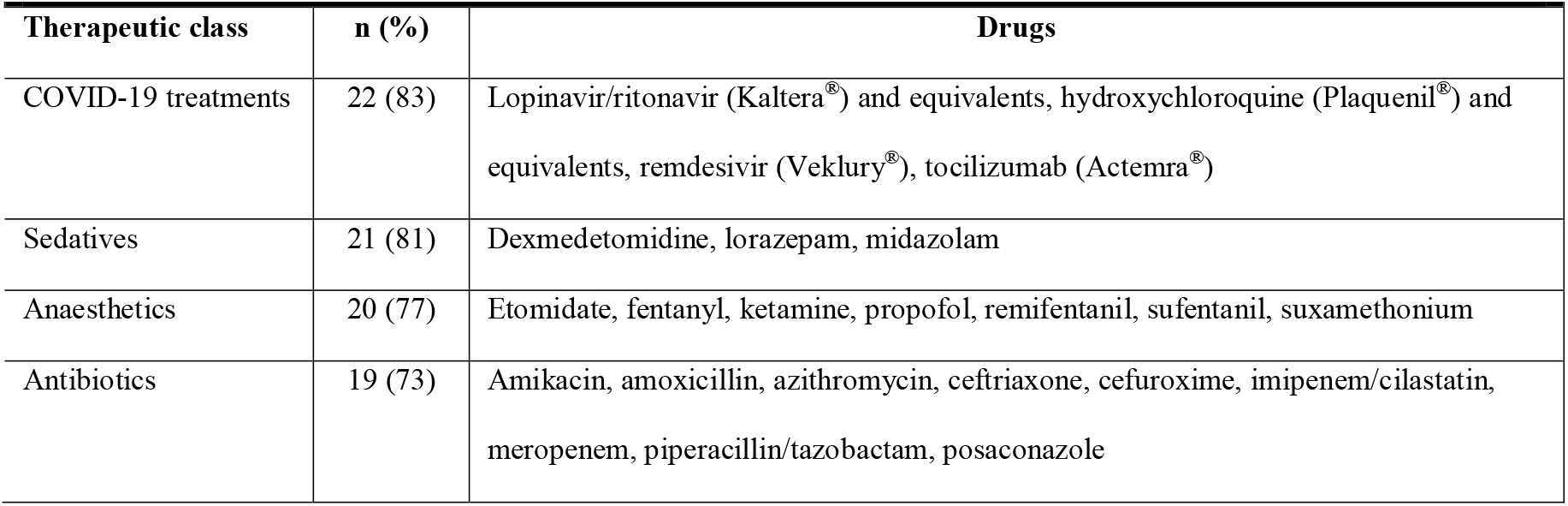
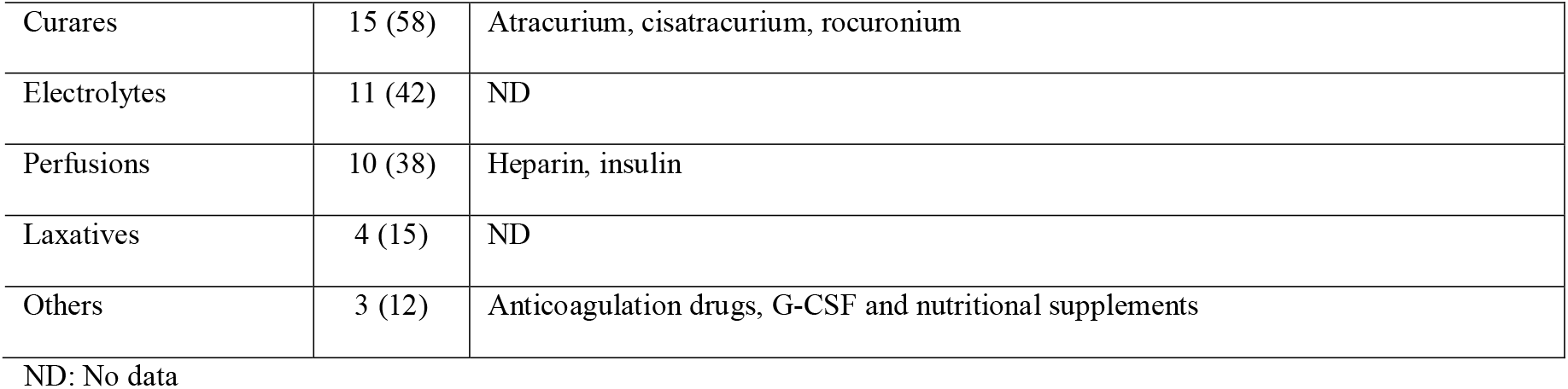
Drugs used for treating COVID-19 patients.

Drug availability in COVID-19 wards was managed by increasing existing stocks (54% of pharmacies; 22/41) and creating extra storage space (51%; 21/41).

Drug production units in 24% of hospital pharmacies (10/41) reconditioned drugs (38% F/I regions, 3/8; 21% G/R regions, 7/33). 10% of hospital pharmacies (4/41) had to produce drugs (25% F/I, 2/8; 6% G/R, 2/33). The main drugs manufactured by hospital pharmacies were hydromorphone, midazolam, morphine, ketamine, fentanyl (all parenteral forms) and hydroxychloroquine (HCQ) suspension.

### SUPPORT FOR MEDICAL AND CARE TEAMS

49% of hospital pharmacies (19/39) worked together with other clinical teams in their hospital to provide appropriate treatment management protocols for COVID-19 patients arriving at the ICU. hospital pharmacies played a key role in many aspects of the COVID-19 pandemic by providing clinical and logistical support to medical and care teams. Detailed support activities for medical and care teams are listed in Table 5.

**Table 5.**
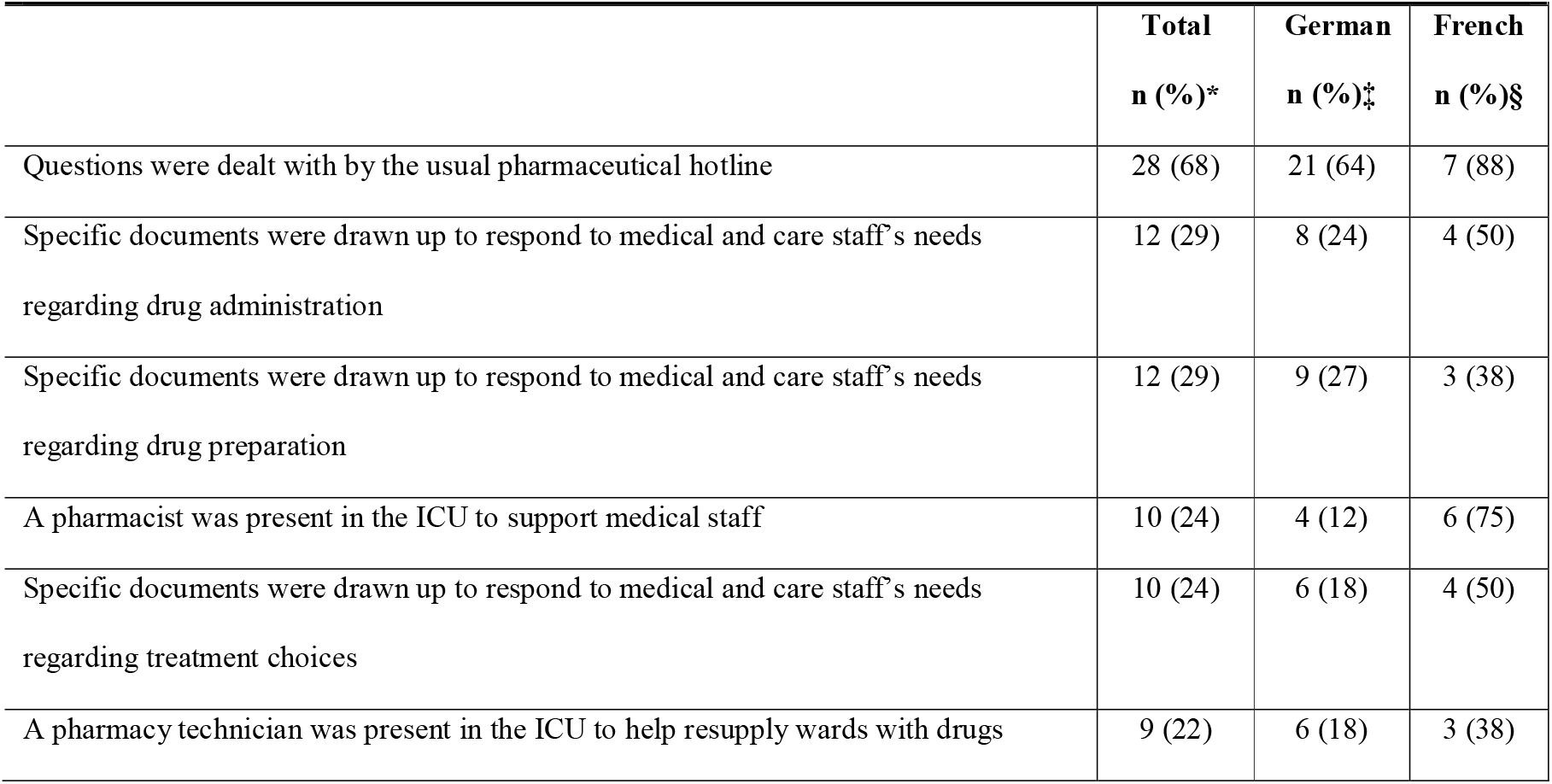
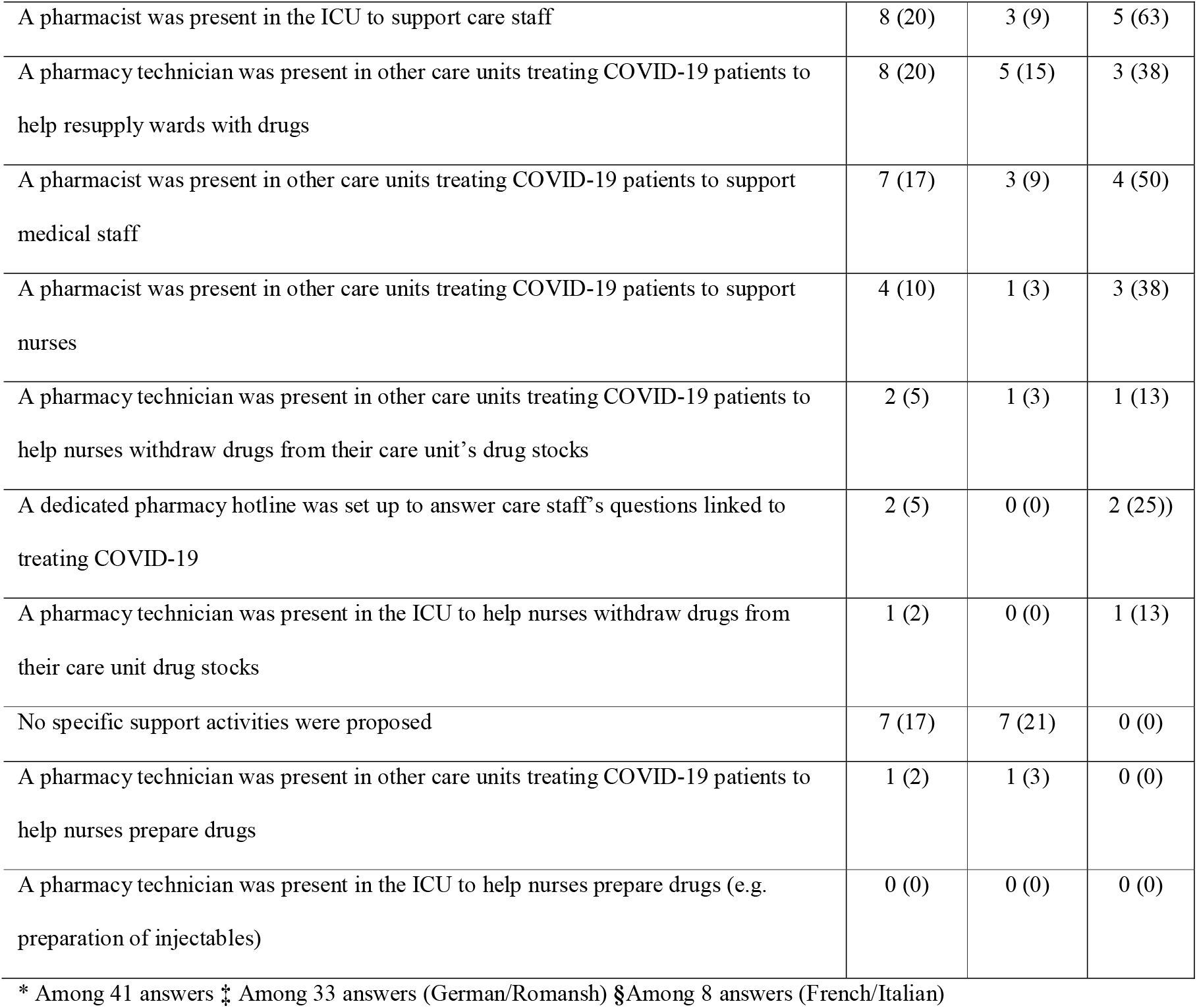
Support activities implemented by hospital pharmacies for medical and care units.

In general, clinical pharmacy services also provided tables with product characteristics and monitored every prescription closely (checking for interactions, contraindications, and correct dosage).

### OTHER ACTIVITIES AND PROBLEMS REPORTED IN WARDS

20% of hospital pharmacies (8/41) implemented other support activities, e.g. a pharmacist and a pharmacy technician were present on weekends to support medical staff (stand-by service at the ICU). They were also in close communication with the ICU head. Pharmacy technicians took over some ICU administrative work or the centralised preparation of prefilled syringes for syringe pumps. Syringes (ketamine, noradrenaline, etc.) for ICUs were produced in hospital pharmacies instead of by nurses. Medication reviews for patient treatments were also carried out. One head hospital pharmacist sat on an interdisciplinary physicians’ committee (emergency, ICU, COVID ward, pneumology, cardiology and infectious diseases). Regarding problems involving the administration of drugs used mainly in ICUs, 12% of hospital pharmacies (5/41) experienced a lack of syringe pumps (25% F/I, 2/8; 9% G/R, 3/33) and 10% (4/41) experienced a lack of injectable drugs (13% F/I, 1/8; 9% G/R, 3/33). Among the hospital pharmacies which faced problems involving the administration of drugs used mainly in ICUs, 60% (9/15) designed alternative drug administration protocols to save on the number of syringe pumps or injectable formulations used, 33% (5/15) managed the distribution and allocation of drug administration material, and 7% (1/15) developed protocols for remdesivir administration.

## DISCUSSION

Most head hospital pharmacists answered our survey, with a repartition in the different regions of Switzerland corresponding relatively to the speaking parts of Switzerland. The French-speaking hospital pharmacies were mainly well prepared. The half of hospital pharmacies moved at-risk employees to safer environments. Moreover, a small part of the participating hospital pharmacies set up a dedicated pandemic response team and provided pharmaceutical support to the medical and nurse’s team in the clinical wards. Some hospital pharmacies assigned a pharmacy team to daily medication stock controls and management in clinical wards treating COVID-19 patients. At the central pharmacy, to limit the risks of shortages, reserve supplies had been planned for in slightly more than half of hospital pharmacies.

Human resource reorganisation was an important issue to guarantee the continuity of the activity during the pandemic. In some countries(21), hospital pharmacies reorganised their activities to limit their human resource risks: roles were redistributed and non-pharmacist staff were used to assist in manufacturing units and answer the phone(22). Other hospital pharmacies encouraged work from home and physical distancing(23). Personal fears regarding contamination risk originating in nursing homes or between colleagues was an issue in some hospital pharmacies and should be considered in any future pandemic preparedness plans.

During crisis management, the presence of a team leader is crucial(24, 25). The pandemic preparedness plan recommends that organisations create a “pandemic team” to manage their response to an outbreak(26). However, in our study, the pandemic’s teams were composed mainly of the head pharmacist and a representative of the pharmaceutical logistics unit (namely a pharmacist in charge of the logistics at hospital pharmacies). A recent hospital pharmacy study, highlighted the difficulty for head pharmacist to lead the crisis management. Furthermore, the leader and his team, must identify quickly the problem and then make decisions, implement management tools, and communicate effectively. For this purpose, dashboards can be helpful (27). In the present survey, among hospital pharmacies with a disaster management plan, the half-created paper/electronic dashboards or other management tools (i.e. an Excel^®^ file) especially for the COVID-19 crisis. The recent study mentioned underlined also the importance of preparing and drill it. Indeed, having management plan is a precious starting point but it is not enough on its own (27). The main point is to test and train it (28). In contrast, fewer hospital pharmacies had prepared a human resource continuity plan. For 18% few a plan existed before the pandemic occurred and for the remaining 18%, the plan was specially created to manage the pandemic: only a small part of German-speaking.hospital pharmacies, but mainly of French-speaking hospital pharmacies. As numbers of contaminations grew during the crisis, a third of French-speaking hospital pharmacies created specific human resource continuity plans, and much more than although only few parts of the German-speaking hospital pharmacies did. This reflected the fact that French-speaking regions suffered more contaminations than German-speaking regions, which thus felt less need to create those plans(29).

Higher incidences of stockouts were to be expected during a pandemic; thus, drug stocks in care units treating COVID-19 patients were managed by either increasing existing stocks or creating extra storage space. Drug management in wards treating COVID-19 patients generated some adjustments to routine roles. At the beginning of the pandemic, it was very difficult for hospital pharmacies to anticipate which drugs would be used the most and thus which had to be ordered, bought, and stocked in sufficient quantities. This raises the question of risk perception, which seems to have been different in French- and German-speaking regions regarding the number of COVID-19 patients expected. It is noteworthy that some of hospital pharmacies experienced no stockouts at all. French-speaking regions made more preventive stocks, but also experienced relatively more stockouts. This might be explained by the fact that French-speaking regions were more affected by the pandemic.

A scoping review of pharmacists’ roles during the pandemic suggested that hospital pharmacies provided drug information to healthcare professionals as part of their daily activities(30). This significant review was the first to highlight that counselling healthcare professionals and patients on drugs were the main action carried out by hospital pharmacists during the COVID-19 pandemic. Indeed, the COVID-19 outbreak revealed new opportunities for hospital pharmacists as they stepped into new roles (greater involvement in thinking about treatment choices, counselling about drugs saving strategy), suggesting that fully integrated, inter-sectoral, inter-professional collaboration is necessary to face crises and public health emergencies effectively(31). Our study highlights the key role played by hospital pharmacies’ in many aspects of the COVID-19 pandemic by providing clinical and logistical support to medical and care teams. Clinical and hospital pharmacists promoted safe, effective medication management among COVID-19 patients and participated in guideline development in partnership with care providers (physicians, infectiologists, and other specialists) and pandemic management experts. Our study found that hospital pharmacists went in clinical wards to support nurses, and hospital pharmacies physicians. The roles of the pharmacists in the wards were varied and included, among others, proposing alternatives to guarantee the continuity of care and save the stocks. In addition, specific COVID-19 documents were drawn up to respond to medical needs with regards to drug administration, preparation, and treatment choices. Indeed, by working together with other clinicians to provide appropriate treatment management protocols for COVID-19 patients arriving at the ICU, pharmacists underline their important role during a pandemic. These roles are supported by international disaster health community who states that pharmacists should have more roles in disasters, in addition to logistics, including stockpile management, vaccination, health and medication education, ensuring continuity of medication care, highlighting the important role for pharmacist in disaster management teams(7, 9).

Future improvements in crisis response will require action at many levels: in hospital pharmacies, hospitals, cantons and nationally. To prevent stock outs, the manufacture of drugs and medical devices within Switzerland should be promoted. Generally, for subsequent waves of the pandemic in Switzerland, the following measures are recommended to hospital pharmacists: 1) update Standard operating procedures and emergency planning procedures; 2) develop worst-case scenario plans for dealing with supply chain and train them (e.g. table top, preparedness exercises etc.); 3) refine business continuity plans; 4) maintain dashboards and concepts used in the first wave (e.g. communication concept); 5) ensure that protection measures and hygiene and social distancing rules are respected; 6) anticipate and ensure minimal pharmacy stock needs by stocking “pandemic inventories” of drugs, disinfectants and Personal Protective Equipment; 7) monitor and evaluate drug use, availability and shortages; 8) exchange directly with stakeholders (wards, public administration, etc.).

In the future, it would be also interesting to repeat this questionnaire to evaluate the response after several waves of COVID-19. Thus, we could measure if the response changes with experience and see if hospital pharmacies having already been strongly impacted by the first wave of the pandemic had an easier response to the next wave.

The present survey had however some limitations. The first is that only 66% head hospital pharmacists who received the questionnaire answered it despite two reminders. Nevertheless, this rate remains high and can be considered representative of pharmacies working in acute hospitals. Pharmacists who were not affiliated to the GSASA were not requested to participate because, as they generally do not work in acute care hospitals (but rather in psychiatric clinics or small private hospitals), they were potentially less affected by the imperatives of COVID-19 crisis management. Also, some participants failed to complete the whole survey, making comparisons between some answers difficult in a few cases.

## CONCLUSIONS

This was the first national-level study on the interventions undertaken by hospital pharmacies in acute care hospitals across Switzerland in response to the vast new challenges encountered during the COVID-19 pandemic. Swiss hospital pharmacies encountered many challenges and had to find solutions quickly, effectively and safely. The survey highlights the key role played by hospital pharmacy’s in many aspects during the pandemic by providing logistical and clinical support to medical and care teams. Altogether, the findings illustrate the wide range of activities undertaken by hospital pharmacies with regards to crisis management, but they identified major areas of concern that emerged due to this public health crisis. The lessons and experiences outlined could be used to improve the quality of the preparation for similar future events in Switzerland such as in any hospital pharmacies in Europe or in other parts of the World.

## Data Availability

Raw data of the survey would be provided upon request

## ABBREVIATION LIST

COVID-19: Coronavirus disease 2019
EpidA: Swiss Federal Act on Epidemics
F: French-speaking region of Switzerland
G: German-speaking region of Switzerland
GSASA: Swiss Society of Public Health Administration and Hospital Pharmacists
I: Italian-speaking region of Switzerland
ICU: Intensive Care Unit
MERS-CoV: Middle East respiratory syndrome coronavirus
R: Romansh-speaking region of Switzerland
SARS-CoV-1: Severe acute respiratory syndrome coronavirus 1
SARS-CoV-2: Severe acute respiratory syndrome coronavirus 2
Swissmedic: Swiss Agency for Therapeutic Products
WHO: World Health Organisation

## DECLARATIONS

### CONSENT FOR PUBLICATION

Not applicable

### AVAILABILITY OF DATA AND MATERIALS

All questions of the survey are available in appendix I. Detailed results of surveys for each question can be asked to the corresponding author.

### CONFLICTS OF INTEREST/COMPETING INTERESTS

The authors have declared no potential conflicts of interest.

### FUNDING AND ACKNOWLEDGMENT

This study was funded by the Swiss Federal Department of Defence, Civil Protection and Sport, through the Centre of Competence for Military and Disaster Medicine. The survey was reviewed by the steering committee of the Swiss Society of Public Health Administration and Hospital Pharmacists, especially PD Dr Johnny Benney, past president.

### CONTRIBUTORSHIP STATEMENT

LS: Conceptualization, Data curation, Formal analysis, Investigation, Methodology, Writing – original draft

YD: Conceptualization, Data curation, Formal analysis, Investigation, Methodology, Writing – original draft

PB: Conceptualization, Funding acquisition, Methodology, Supervision, Writing – review & editing

NW: Conceptualization, Funding acquisition, Methodology, Supervision, Writing – review & editing

## Notes

### Summary of Updates

Improved the general quality of the manuscript and the presentation of the results.

